# Lifestyle associates with unique resistome and microbiome signatures in children

**DOI:** 10.1101/2025.06.25.25330258

**Authors:** Hendrik Dühr, Katariina Pärnänen, Nina Kucháriková, Paulina Werner, Göran Pershagen, Leo Lahti, Harri Alenius, Anna Bergström, Matti O. Ruuskanen, Nanna Fyhrquist

## Abstract

**Background:** Antibiotic resistance is a global health crisis that is not solely explained by antibiotics usage. However, environmental and lifestyle contributions to antimicrobial resistance (AMR) in children are not well understood, especially compared to adults. As the gut functions as a reservoir for antibiotic resistance genes (ARGs), the aim of this study was to better understand the influence of lifestyle on the gut microbiome and resistome using shotgun-metagenomic sequencing data of Swedish children from the PARSIFAL (Prevention of Allergy Risk factors for Sensitization In children related to Farming and Anthroposophic Lifestyle) study.

**Results:** Farm children exhibited high proportions of unique bacterial species and differentially abundant ARGs linked to the farm environment, and similar differences were found in anthroposophic children. Age, breastfeeding duration, and obesity significantly influenced the overall resistance load, independently of lifestyle. Despite limited statistical power, our findings suggest that lifestyle and environment both shape the microbiome and resistome of children.

**Conclusions:** This study corroborates the possible influence of the farm environment on the gut microbiome and resistome, revealing a highly individualized repertoire of low-abundance microbes and ARGs in farm children. Additionally, associations of age, obesity and the duration of exclusive breastfeeding with ARG load were found in a currently understudied age range. Overall, this study raises the need for further research on rare species and ARGs as well as their transmission dynamics in relation to the environment.

## Background

In recent decades antibiotic resistance has become a major global problem affecting human health, currently being estimated to directly cause more than 1 million deaths per year(1), as well as economic costs in the order of billions of dollars, with both numbers expected to rise drastically in the coming decades(2). Especially young children could suffer from potential declines in drug efficacy due to antimicrobial resistance (AMR), as antibiotics currently provide an effective means to prevent child death (3). In this context, the human gut microbiome, i.e., the collection of microbes in the human gut(4), has been increasingly studied as it harbors many antibiotic resistance genes (ARGs), whose entirety make up the gut resistome(5).

Bacteria acquire resistance either through mutations or by horizontal gene transfer (HGT)(6), often mediated by mobile genetic elements (MGEs) such as plasmids, transposons and integrons(7). HGT may facilitate the spread of ARGs between commensal and pathogenic bacteria, making the human gut microbiome an important reservoir for antimicrobial resistance(8).

Even though ARGs occur naturally in the environment and can be traced back to prehistoric ages(9), it has been shown that human activity significantly increases the number of ARGs found in different environments, including the human gut and skin, with 23.78 % of the detected ARGs in all environments being considered a health risk(10). Although antibiotic consumption is a major contributing factor, it does not fully account for the global spread of AMR(11). Furthermore, the transmission dynamics of ARGs between animals, humans and environments are far from being quantified and remain poorly understood(12).

Environmental exposures and lifestyle factors are known to strongly influence the gut microbiome composition(13). Yet most studies on childhood microbiomes and resistomes focus on early infancy(14), leaving a knowledge gap with regards to school-aged children. Lifestyles such as the anthroposophic and farm lifestyle are of particular interest due to distinct characteristics and exposures. The anthroposophic lifestyle is characterized by limited use of antibiotics, antipyretics, and vaccinations, alongside a diet rich in fermented vegetables(15). In contrast, the farm lifestyle has been shown to be protective against allergies, which is potentially caused by a high exposure to microorganisms inhabiting the farm environment, leading to high microbial diversity in the gut(16). However, the exact mechanisms behind this protective effect remain unclear, though current studies suggest a complex interplay between the bacteria and the host immune system(17). In this context, the PARSIFAL study population has been used to investigate protective lifestyle factors for the occurrence of allergic diseases, with results showing that both anthroposophic and farm lifestyles were protective against the occurrence of allergies in children(18,19).

Given AMR’s global threat and the gut microbiome’s role as an ARG reservoir, understanding how childhood lifestyle shapes the resistome is essential. We analyzed metagenomic data from the Swedish participants of the PARSIFAL (Prevention of Allergy Risk factors for Sensitization In children related to Farming and Anthroposophic Lifestyle) study to to assess how anthroposophic and farm lifestyles associate with the gut microbiome and resistome in school-aged children. Our analyses included presence/absence, alpha and beta diversity, as well as differential abundance analyses (DAA), focusing on comparisons between lifestyle and reference groups. Furthermore, we investigated the relation of ARG load (normalized ARG abundance per sample) to lifestyle variables using a generalized linear model (GLM). Additionally, we investigated the relation between ARG load and alpha diversity indices to bacterial class abundances using correlation analysis.

## Materials and methods

### Sample collection

To better understand the lifestyle and environmental drivers of the microbiome and resistome we analyzed 74 stool samples from the cross-sectional PARSIFAL study(18) using paired-end shotgun metagenomic sequencing. The study population consisted of children aged 5-13 years living in the Uppsala and Järna regions (Sweden) and entailed participants living on farms, children with anthroposophic lifestyles and two reference groups of children living in the same areas but not following these lifestyles. For this study, all 74 fecal samples of the Swedish part of the PARSIFAL study were obtained. Samples were taken by the parents, first stored at –20 °C, and finally stored at –80 °C before DNA extraction, as described by Dicksved et al(20). The participant data and samples that were used for the analyses in this study were collected between October 2000 and May 2002. A questionnaire was completed by the parents and has previously been reported in detail by Alfvén et al. (18).

### DNA isolation and quantification

DNA was isolated using the QIAamp® PowerFecal® Pro DNA Kit (Qiagen, Germany, cat. no. 51804) according to manufacturer’s instructions, using the Tissue Lyser II (Qiagen, cat. no. 85300) for lysis. Genomic DNA was eluted in 50 µL of 10 mM Tris. Subsequent quantification and quality control was performed by Novogene (UK) Co., Ltd. using Qubit Fluorometric Quantification (Thermo Fisher, USA).

### Metagenomic sequencing

Library construction was performed according to the following workflow: Genomic DNA was randomly sheared into short fragments that were then end repaired, A-tailed and ligated with Illumina adapters. Fragments with adapters were PCR amplified, size selected, and purified. Paired-end sequencing (read length = 150 base pairs) was performed using the NovaSeq 6000 system (Illumina, USA, cat. no. 20012850)

### Metagenomic analysis

Microbial community profiling of metagenomic sequence reads was performed using MetaPhlAn 4 version 4.0.3(21). We mapped the R1 and R2 reads with Bowtie2 v 2.4.4(22) against the ResFinder database version 2.1.1(23) with the following options: “-D 20 –R 3 –N 1 –L 20 –i S,1,0 5” to identify ARGs. ResFinder was chosen as a database due to its high sensitivity with regards to clinically relevant ARGs such as beta-lactamase encoding genes, compared to other databases(24). The default quality scores in Bowtie were used to ensure high-quality matches are included.

### Data normalization

For taxonomic data, MetaPhlAn relative abundance (%) were transformed into counts through division by 100 and multiplication by library size. Counts were then scaled so that the lowest count > 0 equaled 1. For ARG data, counts were first normalized to gene lengths, and then to library size. As with taxonomic data, counts were scaled so that the lowest count > 0 equaled 1. For the DESeq2 analysis of ARGs, a pseudo-count of 1 was added to all counts of both types of normalized data to allow comparisons between detected and undetected species/genes.

To calculate the ARG load, the raw ARG counts were first normalized to the respective ARG gene length. Then, the sample-wise sums of gene-length normalized ARGs were calculated and normalized to library size. These values were then scaled to represent reads per kilobase of transcript per million mapped reads(25) (RPKM). Library size was chosen for data normalization as it yields similar results compared to 16S rRNA or single marker copy gene normalization(26).

### Rarefaction, alpha and beta diversity

To ensure that the presence/absence analysis was not distorted by differences in groupor library size, we performed additional normalization stepsFirst, MetaPhlAn and ARG counts were normalized to group size before the scaling. Then, rarefaction as defined by Schloss(27), was performed with 100 iterations using rarefy_even_depth from phyloseq 1.50.0(28). The minimum library size was set equal to the smallest sample library size, and subsampling was performed without replacement. The rarefied count tables were then combined to control whether library size affected the presence/absence of species and ARGs. To obtain robust estimates for species alpha and beta diversity measures(27), rarefaction was performed with 100 iterations, using the rarefy_even_depth and getDissimilarity functions of the phyloseq(28) 1.50.0 and mia(29) 1.15.19 packages respectively. As before,the minimum library size was set equal to the smallest sample library size, and subsampling was performed without replacement. Alpha diversities (Shannon(30), inverse Simpson(31), Chao1(32)) were calculated using the estimate_richness function from phyloseq. Beta diversity indices (Bray-Curtis(33), Jaccard(34)) were calculated using the vegdist function (called with getDissimilarity) from the vegan(35) 2.6-8 package. ARG counts were not rarefied before alpha and beta diversity analyses.

### Statistical analysis

Associations between the participant data and the lifestyle groups were analyzed using Fisher’s exact test(36) and the Kruskal-Wallis rank sum test(37), for categorical and continuous variables, respectively. The test statistics were calculated using gtsummary(38) 2.4.0, which was also used to create the final tables. Relative abundances of bacterial classes, families and ARG resistance classes were calculated using the transformAssay function (method = “relabundance”) from the mia(29) 1.15.19 package. The differences in the ratio of the number of unique to shared species and ARGs between the different lifestyle groups were analyzed using a pairwise comparison of proportions implemented in pairwise.prop.test from the stats(39) 4.4.2 package. The venn diagrams were plotted using ggvenn(40) 0.1.10. To test for differentially abundant species between the lifestyle groups and their respective references both MaAsLin2 1.20.0(41) and the Wilcoxon test(42) were used, as previously suggested(43). For both methods, a 10 % prevalence filter was applied. In the Maaslin2 function, log-transformed, TSS-normalized counts were used. As no covariables except the lifestyle groups were used in our analysis, the MAasLin2 results were equal to those of a t-test. As suggested for the identification of differentially abundant genes in metagenomics(44), DESeq2(45) 1.46.0 was used to analyze differentially abundant ARGs between the lifestyle and respective reference groups, using the default settings of the DESeq function. To explore potential relations between the ARG load and the PARSIFAL study variables, a generalized linear model (GLM) was employed, and only participants with complete case records were analyzed (N = 57). The model was built using the variables listed in Table 1. The glm function of the stats(39) package was used to fit the model using the option family “gamma” (link = “log”) as done previously(26). The step function of the stats(39) package was then used to fit the final model, using backwards stepwise regression. Variables were dropped based on the Akaike information criterion. The glht function of the multcomp(46) 1.4-26 was used to perform Tukey’s post hoc test to obtain adjusted p-values for pairwise comparisons. Since our goal was to model ARG load rather than to test associations for specific exposures, we did not define covariates *a priori* but instead started from a full model including all predictors and applied stepwise reduction to obtain the final model. To test potential interplay between bacterial class abundance and ARG load, Shannon(30), inverse Simpson(31) and Chao1(32) indices, correlation analysis was performed. Therefore, classes with less than 50 % prevalence were aggregated to “Other”. Then, transformAssay of the mia(29) package (method = “clr”) was used to CLR transform the counts after adding a pseudocount (half of the minimum positive value of the class counts; 1.5 for bacterial classes, 0.5 for ARG classes). Kendall’s rank correlation was then performed using getCrossAssociation from the mia(29) package. Differences in relative abundances of bacterial classes between the lifestyle and reference groups were calculated and graphed using metacoder(47) 0.3.8. To test differences between lifestyle and reference groups regarding alpha diversities, the Mann-Whitney U Test(48) was performed, as alpha diversities were not normally distributed, which was tested using the Shapiro-Wilk test(49). Principal coordinates analysis(50) (PCoA) was performed using the cmdscale function in the stats(39) package. Unless otherwise stated, p-values were corrected for multiple testing using the Benjamini–Hochberg procedure(51). Results were considered significant for p ≤ 0.05. Plots were created using ggplot2 3.5.1(52). All analyses were performed in R 4.4.2(39).

**Table 1.** | Data for the PARSIFAL study population by lifestyle group. The BMI classification as defined by Cole et al.(53) was used. BMI: Body mass index. Farm: Farm group, Farm ref: Farm reference group, Steiner: Steiner group, Steiner ref: Steiner reference group.

| Variable | Lifestyle group |  |  |  |  | p-value | q-value<sup>2</sup> |
| --- | --- | --- | --- | --- | --- | --- | --- |
|  | Overall<br>N = 74<sup>1</sup> | Farm<br>N = 37<sup>1</sup> | Farm ref<br>N = 11<sup>1</sup> | Steiner<br>N = 19<sup>1</sup> | Steiner ref<br>N = 7<sup>1</sup> |  |  |
| **Sex (male)** | 42 [57%] | 22 [59%] | 7 [64%] | 9 [47%] | 4 [57%] | 0.81<sup>3</sup> | 0.81 |
| **Maternal smoking during pregnancy** | 7 [9.5%] | 5 [14%] | 0 [0%] | 1 [5.3%] | 1 [14%] | 0.52<sup>3</sup> | 0.62 |
| **Parental education** |  |  |  |  |  | <0.001<sup>3</sup> | <0.001 |
| Elementary school or lower | 6 [8.3%] | 5 [14%] | 1 [9.1%] | 0 [0%] | 0 [0%] |  |  |
| Gymnasium | 34 [47%] | 22 [63%] | 8 [73%] | 3 [16%] | 1 [14%] |  |  |
| University | 32 [44%] | 8 [23%] | 2 [18%] | 16 [84%] | 6 [86%] |  |  |
| Unknown | 2 | 2 | 0 | 0 | 0 |  |  |
| **Number of older siblings** |  |  |  |  |  | 0.45<sup>3</sup> | 0.62 |
| 0 | 18 [24%] | 11 [30%] | 3 [27%] | 3 [16%] | 1 [14%] |  |  |
| 1 | 28 [38%] | 12 [32%] | 6 [55%] | 6 [32%] | 4 [57%] |  |  |
| 2 | 14 [19%] | 6 [16%] | 2 [18%] | 4 [21%] | 2 [29%] |  |  |
| ≥3 | 14 [19%] | 8 [22%] | 0 [0%] | 6 [32%] | 0 [0%] |  |  |
| **Environmental smoking at home** | 9 [12%] | 4 [11%] | 2 [18%] | 3 [16%] | 0 [0%] | 0.75<sup>3</sup> | 0.81 |
| **Housholds pets during the first year of life** | 34 [46%] | 26 [70%] | 2 [18%] | 4 [21%] | 2 [29%] | <0.001<sup>3</sup> | <0.001 |
| **Use of antibiotics** |  |  |  |  |  | 0.064<sup>3</sup> | 0.12 |
| Never use | 8 [12%] | 1 [2.9%] | 1 [11%] | 6 [32%] | 0 [0%] |  |  |
| First use >12 months of life | 30 [43%] | 18 [53%] | 3 [33%] | 7 [37%] | 2 [29%] |  |  |
| First use 0-12 months of life | 31 [45%] | 15 [44%] | 5 [56%] | 6 [32%] | 5 [71%] |  |  |
| Unknown | 5 | 3 | 2 | 0 | 0 |  |  |
| **Use of antipyretics** |  |  |  |  |  | <0.001<sup>3</sup> | <0.001 |
| Never use | 11 [16%] | 0 [0%] | 0 [0%] | 10 [53%] | 1 [14%] |  |  |
| First use >12 months of life | 24 [35%] | 18 [55%] | 3 [30%] | 1 [5.3%] | 2 [29%] |  |  |
| First use 0-12 months of life | 34 [49%] | 15 [45%] | 7 [70%] | 8 [42%] | 4 [57%] |  |  |
| Unknown | 5 | 4 | 1 | 0 | 0 |  |  |
| **Child had measles** | 7 [10%] | 0 [0%] | 0 [0%] | 7 [37%] | 0 [0%] | <0.001<sup>3</sup> | <0.001 |
| Unknown | 6 | 5 | 1 | 0 | 0 |  |  |
| **Consumes organic and biodynamic food** | 20 [27%] | 4 [11%] | 1 [9.1%] | 15 [79%] | 0 [0%] | <0.001<sup>3</sup> | <0.001 |
| Unknown | 1 | 1 | 0 | 0 | 0 |  |  |
| **Age (years)** | 8.69 (1.89) | 9.49 (1.85) | 8.73 (1.79) | 7.63 (1.46) | 7.29 (1.11) | <0.001<sup>4</sup> | 0.002 |
| **Length of (exclusive) breastfeeding** |  |  |  |  |  | 0.27<sup>3</sup> | 0.44 |
| Never | 5 [6.8%] | 3 [8.1%] | 1 [9.1%] | 1 [5.3%] | 0 [0%] |  |  |
| 0-4 months | 37 [50%] | 23 [62%] | 5 [45%] | 6 [32%] | 3 [43%] |  |  |
| ≥5 months | 32 [43%] | 11 [30%] | 5 [45%] | 12 [63%] | 4 [57%] |  |  |
| **BMI classification** |  |  |  |  |  | 0.50<sup>3</sup> | 0.62 |
| Not overweight | 56 [78%] | 23 [66%] | 10 [91%] | 17 [89%] | 6 [86%] |  |  |
| Overweight | 13 [18%] | 9 [26%] | 1 [9.1%] | 2 [11%] | 1 [14%] |  |  |
| Obese | 3 [4.2%] | 3 [8.6%] | 0 [0%] | 0 [0%] | 0 [0%] |  |  |
| Unknown | 2 | 2 | 0 | 0 | 0 |  |  |

<sup>1</sup> n [%]; Mean (SD)
<sup>2</sup> Benjamini & Hochberg correction for multiple testing
<sup>3</sup> Fisher's exact test
<sup>4</sup> Kruskal-Wallis rank sum test

## Results

### Study population

The PARSIFAL cross-sectional study has previously been described in detail by Alfvén et al.(18). The Swedish portion of the study population was recruited in Uppsala and Järna (Sweden) and consists of four distinct lifestyle groups: 1. A farm group of children living on farms (N = 37). 2. A farm reference group, consisting of children living in the farmers’ area, but not growing up on farms (N = 11). 3. A group of anthroposophs (N = 19; recruited at Steiner schools and called ‘Steiner’ throughout the text). 4. A Steiner reference group, consisting of children living in the same area but not following the anthroposophic lifestyle (N = 7). There were statistical differences across the lifestyle groups regarding the parental education level, household pet ownership during the first year of life, the use of antipyretics, previous measle infections, the consumption of organic and biodynamic food, and child age (p ≤ 0.05 after correction).

### Community composition

Overall, differences in relative abundances of the microbial taxa and ARGs between the lifestyle groups and references were comparatively small (Fig. 1, Supplementary Fig. 1). There was a high number of low-abundance, non-characterized bacterial classes, especially within Firmicutes (Supplementary Figure 1). Most of the bacterial classes (Fig. 1a) belonged to the class *Clostridia*, followed by *Actinomycetia* and *Bacteroidia*. On the family level (Fig. 1b), most species belonged to three families (*Lachnospiraceae*, *Bifidobacteriaceae* and *Oscillospiraceae*). A total of 8 bacterial genera made up around 50 % of the otal genera in the study group, with *Bifidobacterium* being the most abundant (Fig. 1c). The overwhelming majority of ARGs were tetracycline resistance genes (Fig. 1d). In addition to tetracycline resistance, resistance to four other classes of antibiotics were common (aminoglycosides, amphenicols, beta-lactams, and macrolide–lincosamide–streptogramin B; Fig. 1d).

**Fig. 1.**
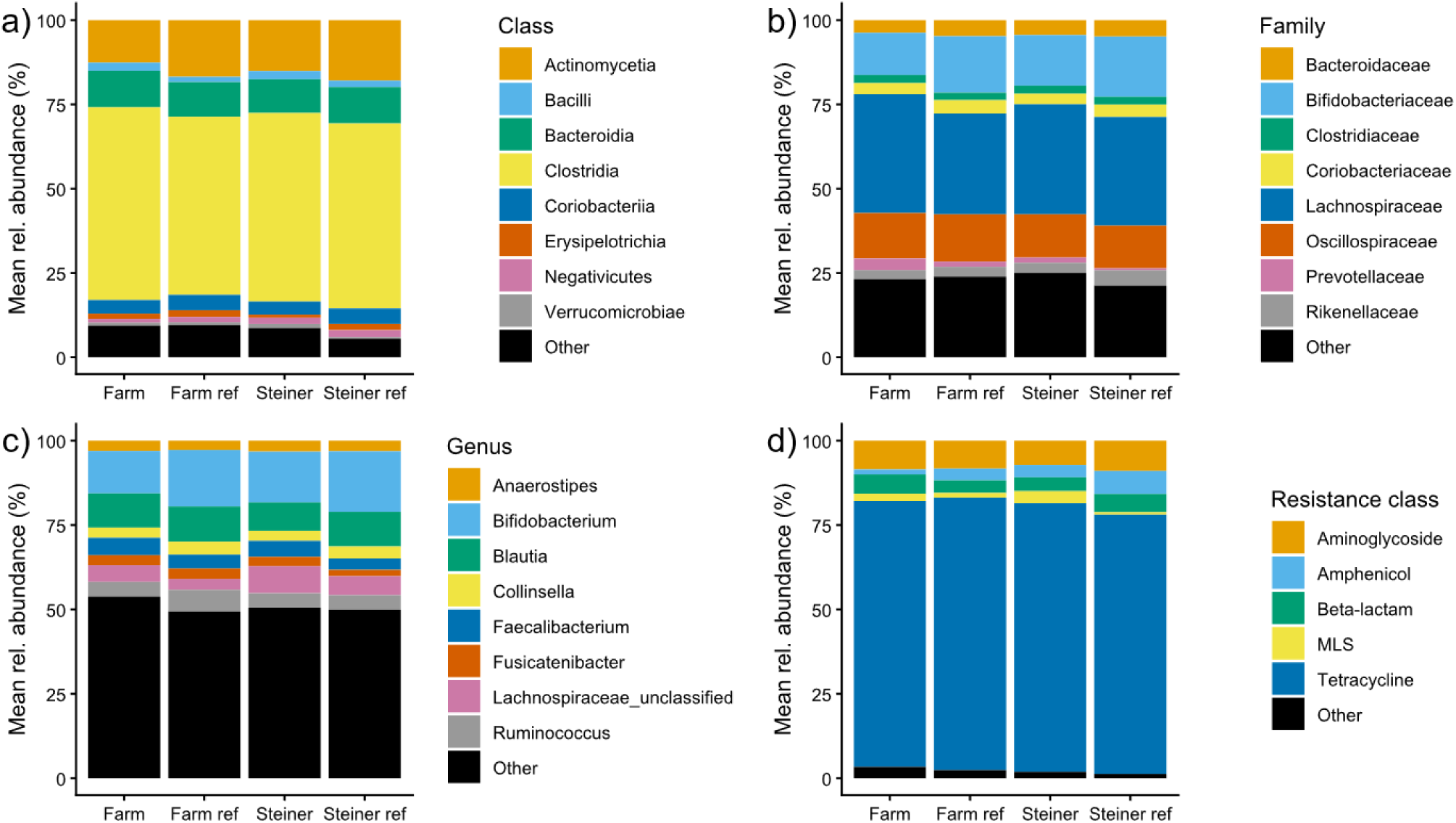
| Microbiome and resistome compositions in the gut of the lifestyle groups. Mean relative abundances (%) are shown on the y-axis. **a** Most abundant bacterial classes. **b** Most abundant bacterial families. **c** Most abundant bacterial genera. **d** Most abundant antibiotic resistance classes. Farm: Farm group (N = 37), Farm ref: Farm reference group (N = 11), Steiner: Steiner group (N = 19), Steiner ref: Steiner reference group (N = 7). MLS: Macrolide, Lincosamide, Streptogramin B.

### Presence/absence analysis

Venn diagrams depicting shared species and ARGs between the lifestyle groups indicated a shared core microbiome and resistome, i.e., shared species and ARGs across all lifestyle groups (Figure 2). Each group had several unique species and ARGs, with the farm and farm reference group containing the highest number of unique species and ARGs. More detailed information regarding the composition of unique species and ARGs is displayed in the supplementary data (Supplementary Figure 2, 3). Additionally, complete lists of all species and ARGs and information on their mean prevalence and relative abundance within the lifestyle groups are provided in Supplementary data 1 and 2.The ratios of unique to shared species were significantly different (p ≤ 0.05) between farmers and all other lifestyle groups, as we l as between Steiner children and both reference groups (Fig. 2b). The ratio of unique/shared ARGs were significantly different (p ≤ 0.05) between the farm group and all other lifestyle groups, though other groups did not significantly differ from each other (Fig. 2d, p > 0.05). The lifestyle exclusive species and ARGs were present mostly in single individuals, as shown in the beta diversity analysis, where no group-dependent groupings were observed (Supplementary Figure 4). Furthermore, more than 90 % of these group-exclusive species and ARGs were of low relative abundance (< 1%) within a given individual. Additionally, we compared the lifestyle and reference groups using alpha diversity indices of species and ARGs but found no significant differences (Supplementary Figure 5, p > 0.05).

**Fig. 2.**
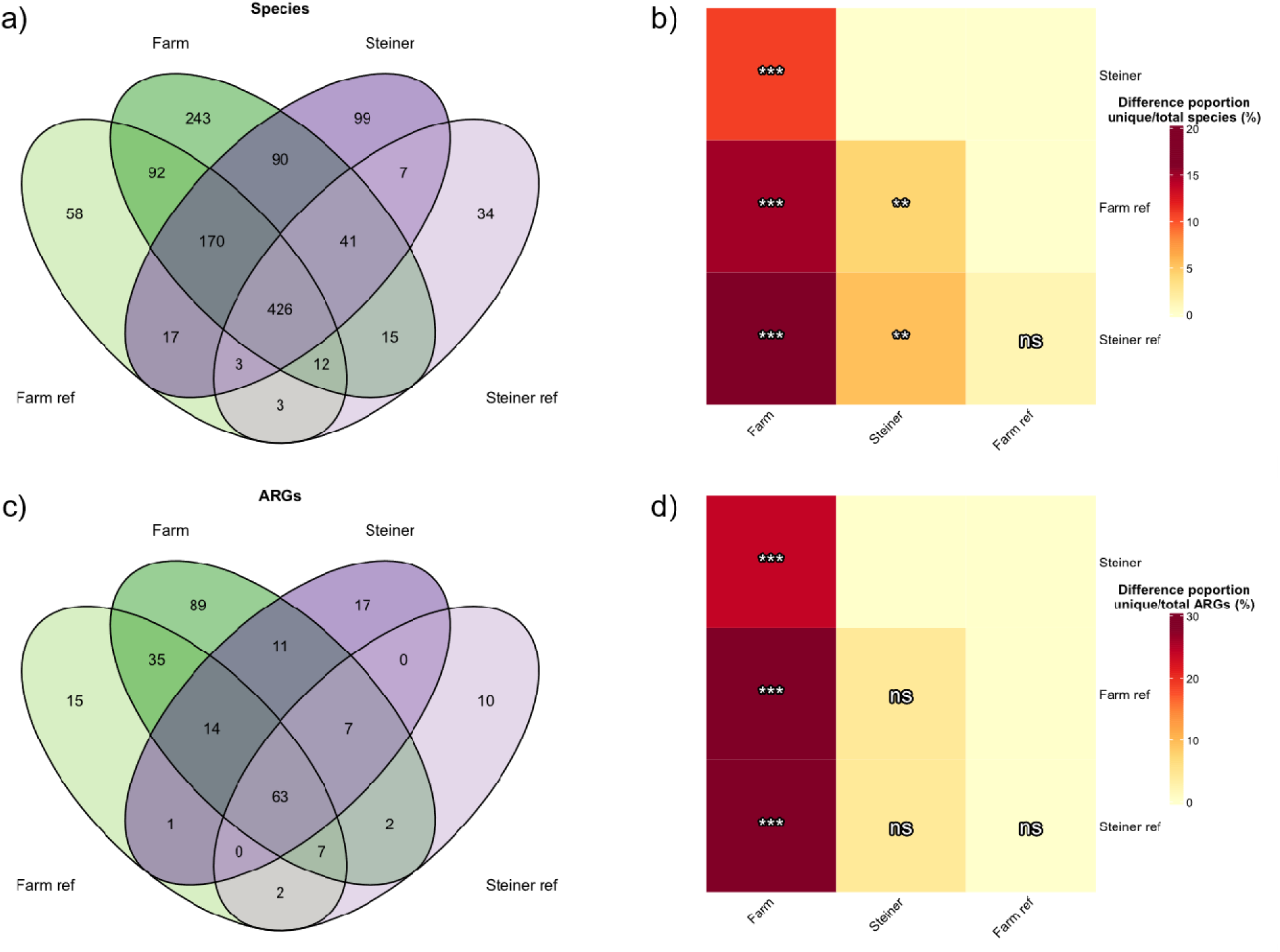
| Venn diagrams and pairwise comparisons of proportions. Venn diagrams depict the shared species (N = 1310) and ARGs (N = 273) between lifestyle and reference groups. **a** presence/absence of species in a lifestyle group, **b** pairwise comparison of proportions of the ratios of unique to non-unique species, **c** presence/absence of ARGs in a lifestyle group, **d** pairwise comparison of proportions of the ratios of unique to non-unique ARGs. Significance of pairwise comparison of proportions is denoted as follows; ns: p > 0.05; *p ≤ 0.05 **p ≤ 0.01; ***p ≤ 0.001. Farm: Farm group (N = 37), Farm ref: Farm reference group (N = 11), Steiner: Steiner group (N = 19), Steiner ref: Steiner reference group (N = 7).

### Differential abundance analyses

To test if any species or ARGs associated with a lifestyle group compared to its reference, differential abundance analysis was performed using MaAsLin2 and the Wilcoxon test for species, and DESeq2 for ARGs. Only one species, *Clostridium sp AM49 4BH* was found to be differentially abundant in farmers compared to the reference group using MaAsLin2 (p ≤ 0.05) but was not found with the Wilcoxon test (p > 0.05). No differential species were detected between Steiner children and their references with either tool (p > 0.05).

Between farmers and references, 19 ARGs were found to be differentially abundant (Fig. 3, p ≤ 0.05), representing 10 of the 22 resistance classes that were detected in the whole study population. Of these, 15 ARGs were differentially abundant in farm children, four in the reference group. *aph(3’’)-Ib* was found to be differentially abundant in Steiner references compared to Steiner children (log2 fold = –6.58, p ≤ 0.05).

**Fig. 3.**
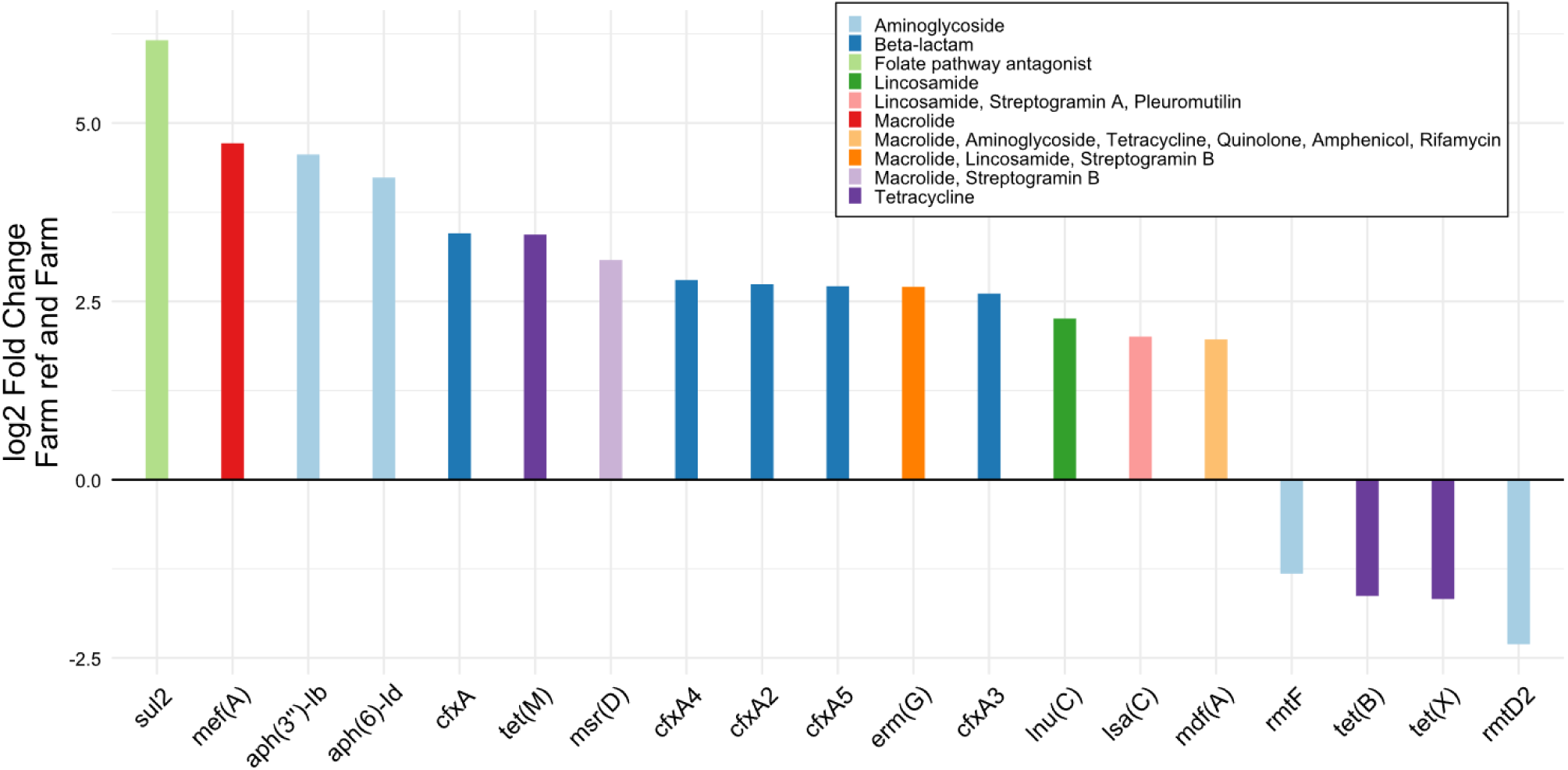
| Differentially abundant ARGs and their corresponding resistance classes between farm children and their reference group. All genes shown were found to be significant in DESeq2 (p ≤ 0.05). The y-axis shows log2 fold changes, the x-axis denotes gene names, positive log2 fold changes indicate differential abundance in the farm group, negative changes indicate differential abundance in the farm reference group. Farm: Farm group (N = 37), Farm ref: Farm reference group (N = 11), Steiner: Steiner group (N = 19), Steiner ref: Steiner reference group (N = 7).

### ARG load

To study the associations of the study variables with the ARG load, a GLM was fitted using stepwise regression, using only complete case records for the variables listed in Table 1 (N = 57, see Supplementary Table 1 for a full description). The body mass index (BMI) category (Fig. 4a), length of exclusive breastfeeding (Fig. 4b) and age (Fig. 4c) were significantly associated with ARG load (p ≤ 0.05). Overweight children had a significantly lower ARG load than non-overweight children (fold change: 0.75, 95 % Confidence interval [CI]: 0.60-0.94). Children breastfed exclusively for more than 5 months had a significantly lower ARG load than children breastfed for shorter periods (fold change: 0.83, 95 % CI: 0.70-0.98). Increasing age (in years) was significantly associated with a lower ARG load (fold change: 0.92, 95 % CI: 0.88-0.97).

**Fig. 4.**
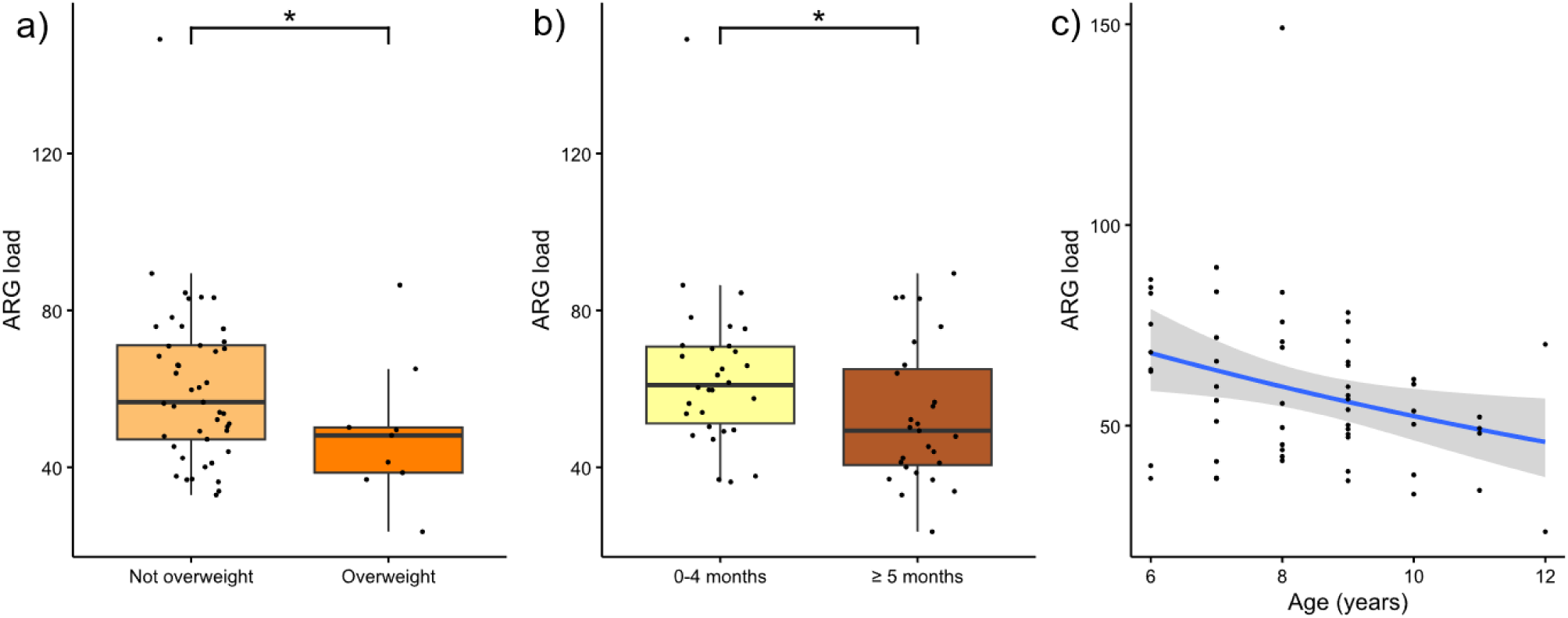
| ARG load in relation to PARSIFAL study variables. To investigate the effects of variables on the ARG load a gamma-distributed GLM model was fitted using backward stepwise-regression. ARG load (N = 57) by: **a** BMI (not overweight and overweight), **b** length of breastfeeding (non-breastfed category not shown) and **c** age (*p* ≤ 0.05). The y axis denotes ARG load (ARG counts normalized to gene lengths and library size, scaled to RPKM). Significance levels are denoted as follows (Tukey’s post-hoc test): *p ≤ 0.05. Boxplot hinges represent 25 % and 75 % percentiles; the center line represents the median. The length of whiskers denotes 1.5 × IQR. The x-axis in c represents age in years. A regression line was fitted using gamma-distributed GLMs. Not overweight: Non-overweight children (N = 45), Overweight: Overweight children(53). 0-4 months: exclusive breastfeeding for less than 5 months (N = 30), ≥ 5 months: exclusive breastfeeding for 5 or more months (N = 27).

### Correlation analysis

We investigated associations between ARG load and alpha diversity with bacterial and ARG class abundance using correlation analysis, as both ARG load and alpha diversity have been linked to human health. The abundances were center log ratio (CLR) transformed to remove compositionality bias. The ARG load and alpha diversity were not significantly correlated with the CLR transformed abundances of ARG classes (all p > 0.05; Supplementary Figure 6). Conversely, we found significant correlations between ARG load and alpha diversity indices and the CLR transformed abundances of bacterial classes (Fig. 5). Overall, the different alpha diversity indices showed similar correlation patterns regardless of being based on relative abundance of species/ARGs (Shannon, inverse Simpson), or estimating the total number of species based on presence/absence data and rare taxa (Chao1). Several unclassified bacterial classes correlated positively with alpha diversity (p ≤ 0.05); *Actinomycetia*, *Clostridia*, *Erysipelotrichia*, and *Coriobacteria* were strongly negatively correlated with all alpha diversity indices. In contrast, *Verrucomicrobiae*, *Eryisipelotrichia* and *Actinomycetia* were the only classes to significantly, though weakly, correlate with the ARG load, with the latter two correlating negatively and *Verrucomicrobiae* correlating positively with the ARG load (Fig. 5).

**Fig. 5.**
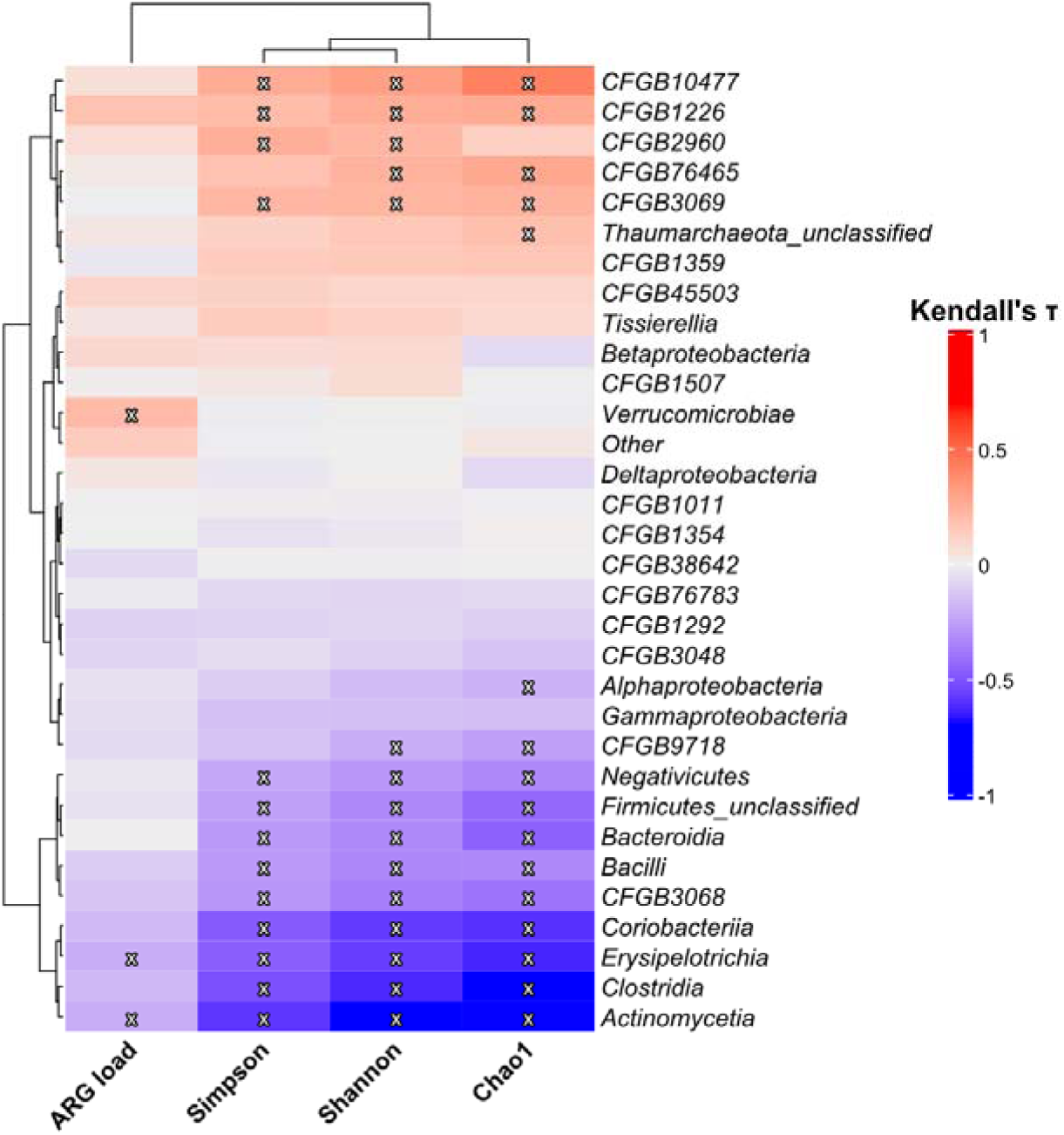
| Correlation of bacterial taxa with ARG load and inverse Simpson, Shannon and Chao1 diversities. CLR transformed bacterial class abundances were correlated with diversity measures using the Kendall rank correlation coefficient (τ). Significant correlations (BH corrected p ≤ 0.05) are denoted with an x. Dendrograms are the result of row– and column-wise hierarchical clustering (complete-linkage) using Euclidian distances. Classes with a prevalence < 50% were aggregated to “Other”. Bacterial class names according to Genome Taxonomy Database (GTDB)(54).

## Discussion

Our study revealed differences in species and ARG presence and absence between the groups, and identified differentially abundant ARGs in the farm group. Furthermore, we were able to identify and corroborate specific lifestyle factors that were associated with variation in ARG load in school-aged children. Nonetheless, these results should be read carefully in the light of the comparatively small sample size and unequal group sizes.

The presence/absence analysis showed that differences in the gut microbiome of both farm and anthroposophic children were primarily driven by the presence of rare, low-abundance species, with both farm and Steiner children having significantly higher proportions of group-exclusive species compared to the reference groups. Interestingly, farm children also had a significantly higher proportion of group-exclusive ARGs compared to all other lifestyle groups. As shown by the non-existent grouping in our beta diversity analysis, which included the presence/absence based Jaccard index, these group-exclusive species and ARGs are highly individual. These findings align with previous evidence of a strong impact of the farm environment on the human gut microbiome and resistome(55), supported by the presence of multiple farm-exclusive species that were strongly associated with farm animals, such as *Mogibacterium kristiansenii, Baileyella intestinalis*(56) (both pigs), *Candidatus Neoanaerotignum tabaqchaliae*(57) (chicken) and *Clostridium cuniculi*(58) (rabbits). In anthroposophic children group-exclusive species included bacteria associated with fermentation of foods (*Leuconostoc citreum*(59), *Lactobacillus farciminis*(59)) and/or are lactic acid bacteria (*Pediococcus pentosaceus*(60), *Ligilactobacillus salivarius*(61)). Lactic acid bacteria have long been discussed to have a variety of beneficial health effects(62), including on the immune system, and may be present in this lifestyle group due to their higher consumption of fermented foods. However, additional studies are needed to determine the persistence of these species. Previous studies focused on the association between the anthroposophic lifestyle and the microbiome identified certain factors (e.g., low antibiotic use, opting for home birth) related to this lifestyle that might affect the gut microbial composition in infants and children(15,63). However, these studies cannot be directly compared to our study due to differences in age (subjects below 2 years of age), methodology and study design. Given the small sample sizes in the Steiner and respective reference groups in this study, differences associated with the anthroposophic lifestyle that were found in the aforementioned studies, e.g., higher abundances of *Bifidobacterium* and *Lactobacilli* might become discernible with increased sample sizes.

Overall, relative abundances of bacterial classes and families observed in the PARSIFAL study closely resemble those found in adult fecal microbiomes(64). While the microbial composition on class and family levels indicated slight variations at higher taxonomic levels between the groups, these differences were non-significant and could not be found with differential abundance analysis. Furthermore, our analysis of alpha diversities did not confirm the differences in alpha diversity that were previously detected in the PARSIFAL study population(20). The discrepancies between the original study and our study may be due to differences in methodology, as our metagenomic analysis was untargeted compared to the targeted T-RLFP method employed in the original study. Furthermore, it should be mentioned that the original 2007 study used a larger subsample from several European countries, which may influence comparisons. The whole PARSIFAL study group will need to be analyzed to resolve the different findings of the studies.

Our study lacks the statistical power to draw definitive conclusions about the protective effects of farm and anthroposophic lifestyles on respiratory allergies which have previously been observed in the original study(18). Nonetheless, our findings are in line with the biodiversity hypothesis(65), particularly given the high number of unique species in the farm group. Considering the well-established role of the (gut) microbiome in shaping immune function(66,67) the presence of a high number of group-exclusive taxa could merit further investigation. Expanding the analysis to include the full study group and/or future studies might help clarify the associations between the environmental exposure to microbes, the gut microbiome and protection against allergic diseases in humans(68).Given the strong protective influence of soil(69) and dust(70) exposure on the microbiome and immune function, the inclusion environmental measurements will be essential in future research.

In contrast to our presence/absence analysis of bacterial species, we only found one species, *Clostridium sp AM49 4BH*, to be differentially abundant in farm children compared to their references. This species has not been associated with the farm lifestyle yet, but has been associated with various diseases, including IBD(71), kidney stone formation(72) and postural orthostatic tachycardia syndrome(73). This result is not surprising, as we employed the commonly used prevalence filter cutoff of 10 %, which removes rare and often low-abundance taxa from DAA analyses(74). However, focusing solely on DAA to analyze differences between groups omits differences regarding rare species that might be part of the core microbiome(75). Though environment specific origins of these rare taxa are likely, we are unable to infer from our data whether the group-specific species are persistent or not, raising the need for studies that include multiple points of sampling.

The differentially abundant ARGs we identified in the farm group compared to the reference group contain numerous ARGs that have been identified in the farm environment, including *sul2*, *mef(A)*, *tetM* and *lnu(C)*, whilst others such as *aph(3*′′*)-IIb* or *msr(D)* have been identified in farm-related seeding sludges (76). This may indicate the farm environment as a potential risk factor for the acquisition of antibiotic resistance genes. The transmission of species that carry ARGs could occur even airborne, as Bai et al.(77) found that this transmission route could be relevant even at a distance from the actual farm environment and thus may not require direct contact with ARG carriers such as farm animals. However, our study cannot quantify the specific contribution of the farm environment to ARG acquisition, as it lacks environmental samples.

In this study, higher age was associated with decreased ARG loads. Increased age has previously been associated with a reduced ARG load in the human gut, demonstrated in a comparative study between infants (age 11-24 months) and young adults (age 17-21)(78). Furthermore, the results of the current study could indicate that extended and exclusive breastfeeding may be beneficial in reducing the overall ARG load, though longitudinal studies are needed to explore this association. In general, breastfeeding has been identified as a factor that lowers the ARG load in infants compared to formula-based diets(26). However, it remains unclear to what extent this effect is due to a reduced exposure to antibiotic resistant bacteria in dietary components other than breastmilk, or due to molecular effects driven by breastmilk consumption(79). Additionally, our study found a lower ARG load to be associated with overweight children compared to non-overweight children. This result aligns with observations in a large cohort from Finland(80) that found an inverse association of BMI and ARG load in adults and pointed to the potential lack of ARG containing foods (fresh vegetables and poultry) in high-fat and high-processed foods and diets. However, these findings are purely observational, and due to the lack of detailed dietary data, our study is unable to explore such associations further. Nonetheless, given the association of high phylogenetic diversity of foods with a low ARG load in adults(81) further investigations are warranted, especially since a phylogenetically diverse diet is generally associated with a healthy lifestyle(82).

*Verrucomicrobiae* was the only bacterial class whose abundance was positively correlated with the ARG load. This may be due to a high competitiveness of the *Verrucomicrobia* phylum after antibiotic treatment(83), indicating a potential competitive advantage of this class in high ARG load environments. However, this may also be due to the fact that the host range of ARGs is taxonomically limited(84). The negative correlations between ARG load and bacterial classes (*Erysipelotrichia* and *Actinomycetia*) were few and comparatively weak in the current study. To the best of our knowledge, literature on these associations is currently sparse.

A lowered abundance of *Erysipelotrichia* after antibiotic treatment has been reported in gnotobiotic mice(85), contrasting with our findings in humans. The negative correlation between *Actinomycetia* abundance and ARG load is unexpected, given its known production of antimicrobial compounds(86). However, our analysis cannot determine causality. The many significant correlations between bacterial class abundances and alpha diversity compared to ARG load suggest a decoupling of species diversity and ARG load. However, further analyses with larger study groups are needed to identify the bacterial drivers of ARG load in children as demonstrated previously in infants (87).

Many current studies examining the microbiome and resistome in the gut of children focus on the first few weeks to months after birth, considering specific factors such as nutrition and antibiotic treatment that enable well-defined study groups(26,88). Additionally, not all these studies take both the microbiome and resistome into account in their analyses, and only cover a maximum age of 5 years(14). In contrast, this study examined both the microbiome and resistome in distinct lifestyle groups (farmers, anthroposophs) with an age range (5–13) that is not adequately represented in the current literature. Nonetheless, larger sample sizes and longitudinal studies that focus on the farm environment are needed to explore connections between the environment, microbiome and resistome, as our study lacks statistical power and a balanced design with similar group sizes.

## Conclusions

In conclusion, high-abundance taxa and ARGs were largely similar across groups, while the farm environment appears to influence the gut microbiome and resistome primarily through low-abundance, highly individualized species and ARGs. We also observed associations between lifestyle variables (age, obesity, length of exclusive breastfeeding) and ARG load in a currently understudied age range. These findings highlight the potential role of environmental and lifestyle factors in shaping the resistome and support further research focusing on rare taxa, ARGs and their transmission dynamics.

## Declarations

### Ethics approval and consent to participate

This study was approved by Karolinska Institutet’s regional research ethics committee. Informed consent was obtained from the parents of each child for the questionnaire and the stool sample (Dnr 02-475 and 00-140).

### Consent for publication

Not applicable.

### Availability of data and materials

The datasets analyzed in this study are not publicly available due to the private and sensitive nature of the research data, which could compromise participant privacy. However, the data can be made available by the corresponding authors upon reasonable and qualified request.

### Availability of computer code and software

The analysis code for this study is available from the GitHub repository Duehr_et_al_2025_supplementary_code and can be accessed via the following link: https://doi.org/10.5281/zenodo.15534317.

### Authors’ contributions

H.D.: Data analysis, Visualization, Writing – Original Draft, Review, Editing. K.P. & M.O.R.: Conceptualization, Supervision, Metagenomic data analysis, Writing – Review, Editing. N.K. & P.W.: DNA isolation, Writing – Review, Editing. G.P. & A.B.: Cohort data curation, Resources, Writing – Review, Editing. L.L. & H.A.: Resources, Funding, Writing – Review, Editing. N.F.: Conceptualization, Supervision, Resources, Funding, Writing – Review, Editing. All authors approved the final version.

## Data Availability

https://doi.org/10.5281/zenodo.15534317

## Acknowledgments

We wish to acknowledge CSC – IT Center for Science, Finland, for computational resources. We thank the children and parents for participating in the PARSIFAL study. We also thank all the staff involved in the study.

