## Supplements for "Lifestyle associates with unique resistome and microbiome signatures in children"

### Supplementary information

**Supplementary Data 1 | Species prevalence and abundance within the different lifestyle groups.** prevalence_Farm: Species prevalence in the farm group. prevalence_Farm_ref: Species prevalence in the farm reference group. prevalence_Steiner: Species prevalence in the Steiner group. prevalence_Steiner_ref: Species prevalence in the Steiner reference group. relabundance_Farm: Mean species relative abundance in the farm group. relabundance_Farm_ref: Mean species relative abundance in the farm reference group. relabundance_Steiner: Mean species relative abundance in the Steiner group. relabundance_Steiner_ref: Mean species relative abundance in the Steiner reference group. The data is on a scale from 0-1.

**Supplementary Data 2 | ARG prevalence and abundance within the different lifestyle groups.** prevalence_Farm: ARG prevalence in the farm group. prevalence_Farm_ref: ARG prevalence in the farm reference group. prevalence_Steiner: ARG prevalence in the Steiner group. prevalence_Steiner_ref: ARG prevalence in the Steiner reference group. relabundance_Farm: Mean ARG relative abundance in the farm group. relabundance_Farm_ref: Mean ARG relative abundance in the farm reference group. relabundance_Steiner: Mean ARG relative abundance in the Steiner group. relabundance_Steiner_ref: Mean ARG relative abundance in the Steiner reference group. The data is on a scale from 0-1.

**Supplementary Table 1 | Participant data used in the generalized linear model.** This table contains only data for study participants whose records were complete for all the listed variables. SD: Standard deviation, q-value: p-value corrected for multiple testing. BMI: Body mass index. Farm: Farm group, Farm ref: Farm reference group, Steiner: Steiner group, Steiner ref: Steiner reference group.

| | | Lifestyle group | | | | | |
|---|---|---|---|---|---|---|---|
| **Variable** | **Overall** N = 57[1] | **Farm** N = 23[1] | **Farm ref** N = 9[1] | **Steiner** N = 18[1] | **Steiner ref** N = 7[1] | **p-value** | **q-value**[2] |
| **Sex (male)** | 31 [54%] | 12 [52%] | 6 [67%] | 9 [50%] | 4 [57%] | 0.92[3] | 0.92 |
| **Maternal smoking during pregnancy** | 4 [7.0%] | 2 [8.7%] | 0 [0%] | 1 [5.6%] | 1 [14%] | 0.80[3] | 0.86 |
| **Parental education** | | | | | | <0.001[3] | 0.002 |
| Elementary school or lower | 3 [5.3%] | 2 [8.7%] | 1 [11%] | 0 [0%] | 0 [0%] | | |
| Gymnasium | 24 [42%] | 13 [57%] | 7 [78%] | 3 [17%] | 1 [14%] | | |
| University | 30 [53%] | 8 [35%] | 1 [11%] | 15 [83%] | 6 [86%] | | |
| **Number of older siblings** | | | | | | 0.31[3] | 0.44 |
| 0 | 16 [28%] | 9 [39%] | 3 [33%] | 3 [17%] | 1 [14%] | | |
| 1 | 21 [37%] | 7 [30%] | 5 [56%] | 5 [28%] | 4 [57%] | | |
| 2 | 10 [18%] | 3 [13%] | 1 [11%] | 4 [22%] | 2 [29%] | | |
| ≥3 | 10 [18%] | 4 [17%] | 0 [0%] | 6 [33%] | 0 [0%] | | |
| **Environmental smoking at home** | 5 [8.8%] | 1 [4.3%] | 1 [11%] | 3 [17%] | 0 [0%] | 0.49[3] | 0.64 |
| **Housholds pets during the first year of life** | 24 [42%] | 17 [74%] | 1 [11%] | 4 [22%] | 2 [29%] | <0.001[3] | 0.002 |
| **Use of antibiotics** | | | | | | 0.041[3] | 0.077 |
| Never use | 7 [12%] | 0 [0%] | 1 [11%] | 6 [33%] | 0 [0%] | | |
| First use >12 months of life | 24 [42%] | 13 [57%] | 3 [33%] | 6 [33%] | 2 [29%] | | |
| First use 0-12 months of life | 26 [46%] | 10 [43%] | 5 [56%] | 6 [33%] | 5 [71%] | | |
| **Use of antipyretics** | | | | | | <0.001[3] | <0.001 |
| Never use | 11 [19%] | 0 [0%] | 0 [0%] | 10 [56%] | 1 [14%] | | |
| First use >12 months of life | 17 [30%] | 11 [48%] | 3 [33%] | 1 [5.6%] | 2 [29%] | | |
| First use 0-12 months of life | 29 [51%] | 12 [52%] | 6 [67%] | 7 [39%] | 4 [57%] | | |
| **Child had measles** | 7 [12%] | 0 [0%] | 0 [0%] | 7 [39%] | 0 [0%] | <0.001[3] | 0.002 |
| **Consumes organic and biodynamic food** | 15 [26%] | 1 [4.3%] | 0 [0%] | 14 [78%] | 0 [0%] | <0.001[3] | <0.001 |
| **Age (years)** | 8.33 (1.62) | 8.96 (1.80) | 8.67 (1.32) | 7.78 (1.35) | 7.29 (1.11) | 0.029[4] | 0.063 |
| **Exclusive breastfeeding ≥5 months** | 27 [47%] | 6 [26%] | 5 [56%] | 12 [67%] | 4 [57%] | 0.059[3] | 0.10 |
| **BMI classification** | | | | | | 0.60[3] | 0.71 |
| Not overweight | 45 [79%] | 15 [65%] | 8 [89%] | 16 [89%] | 6 [86%] | | |
| Overweight | 9 [16%] | 5 [22%] | 1 [11%] | 2 [11%] | 1 [14%] | | |
| Obese | 3 [5.3%] | 3 [13%] | 0 [0%] | 0 [0%] | 0 [0%] | | |

[1] n [%]; Mean (SD)
[2] Benjamini & Hochberg correction for multiple testing
[3] Fisher's exact test
[4] Kruskal-Wallis rank sum test

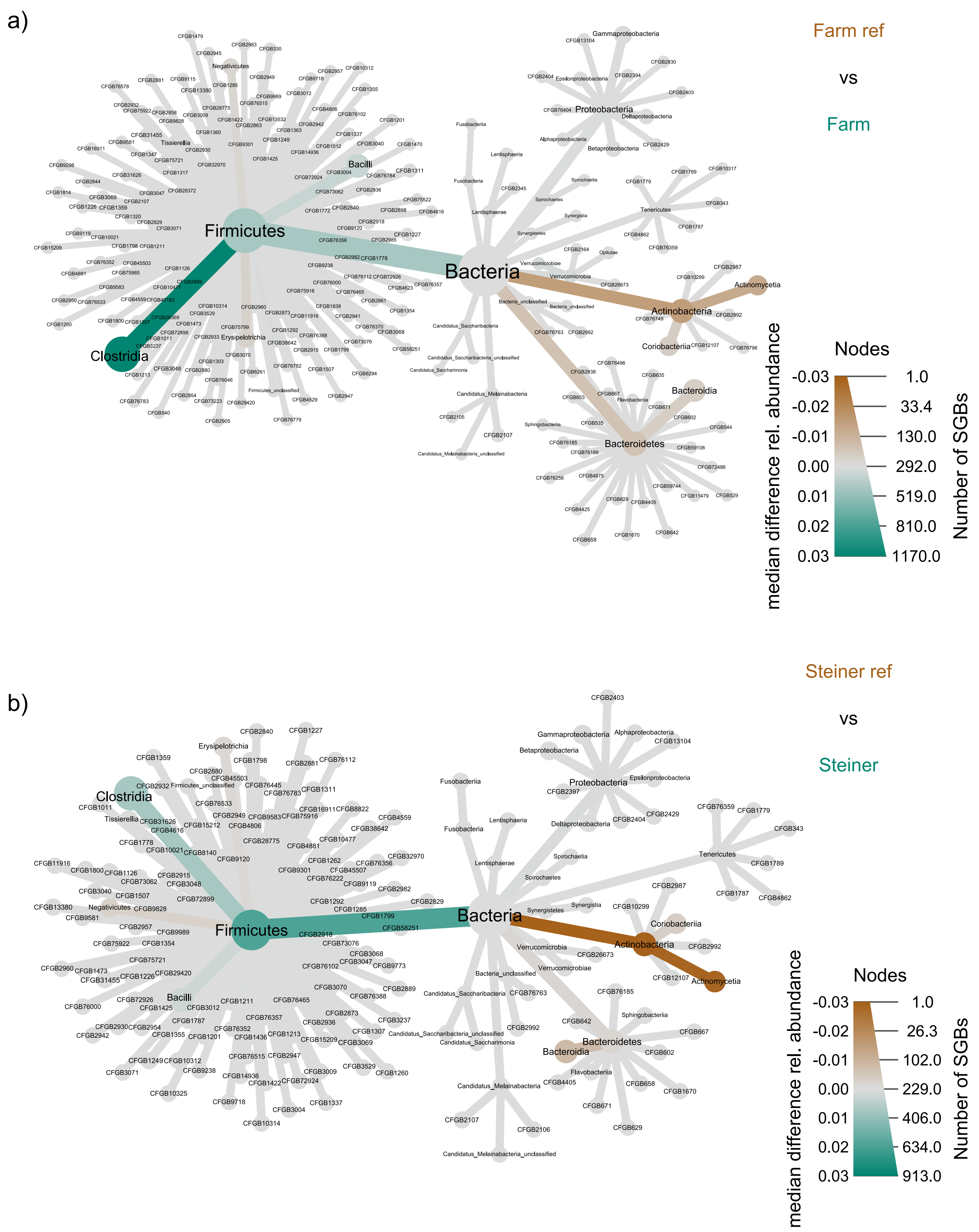


**Supplementary Figure 1 | Differences in relative abundances between lifestyle and reference groups.** Brown-tones indicate higher median relative abundance in the reference groups, green-tones higher abundance in the lifestyle groups. The node sizes show the number of species-level genome bins (SGBs) within a phylogenetic level. **a** Differences between farm references (N = 11) and farm children (N = 37). **b** Differences between Steiner references (N = 7) and Steiner children (N = 19).

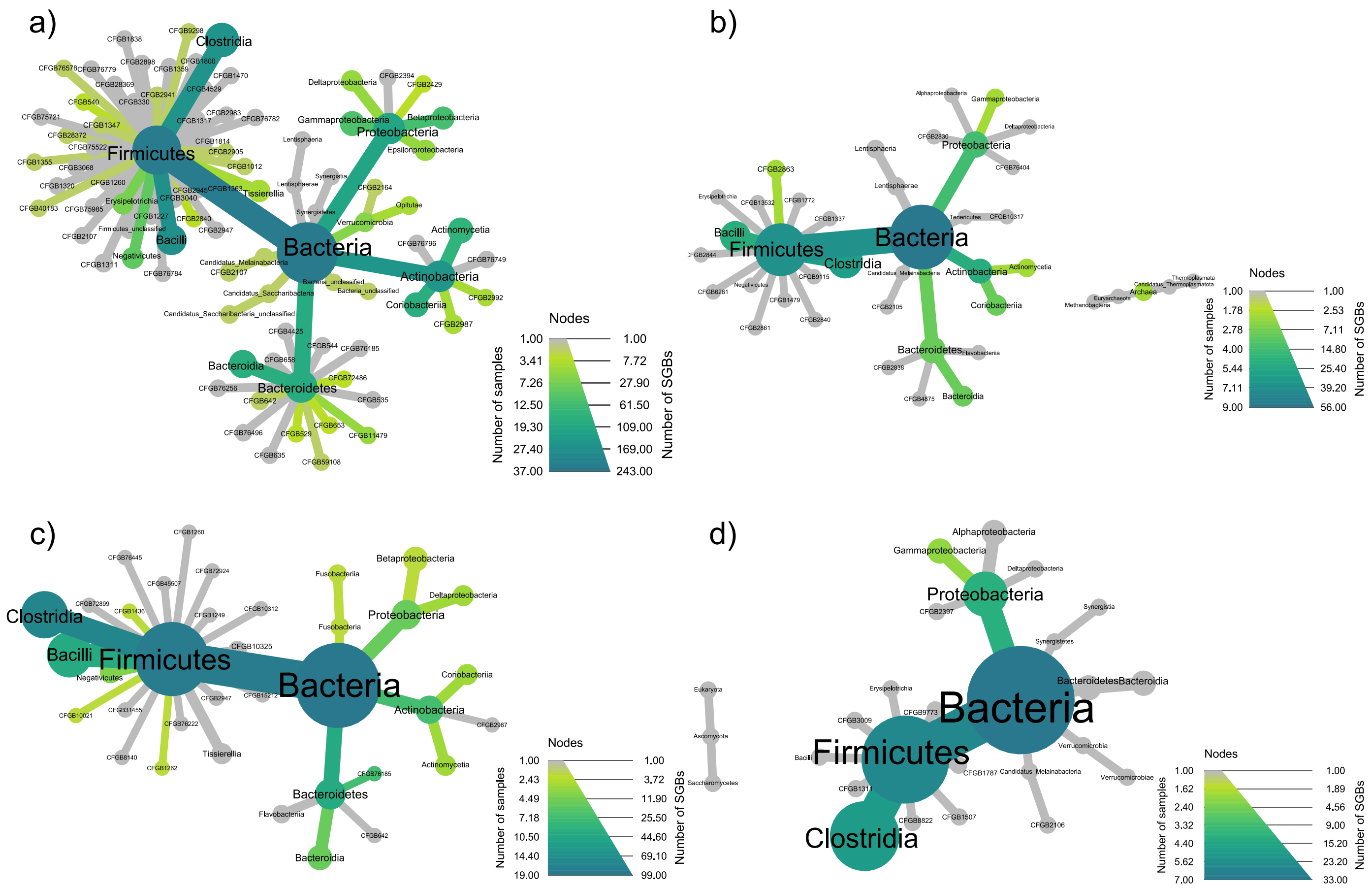


**Supplementary Figure 2 | Taxonomic composition of group-exclusive species.** Shown is the phylogenetic composition of the group exclusive species for **a** farm children (N = 37), **b** farm references (N = 11), **c** Steiner children (N = 19) and **d** Steiner references (N = 7). The colour scheme refers to the number of samples a given taxonomic unit is found in, the node size refers to the number of species-level genome bins (SGBs) within a phylogenetic level.

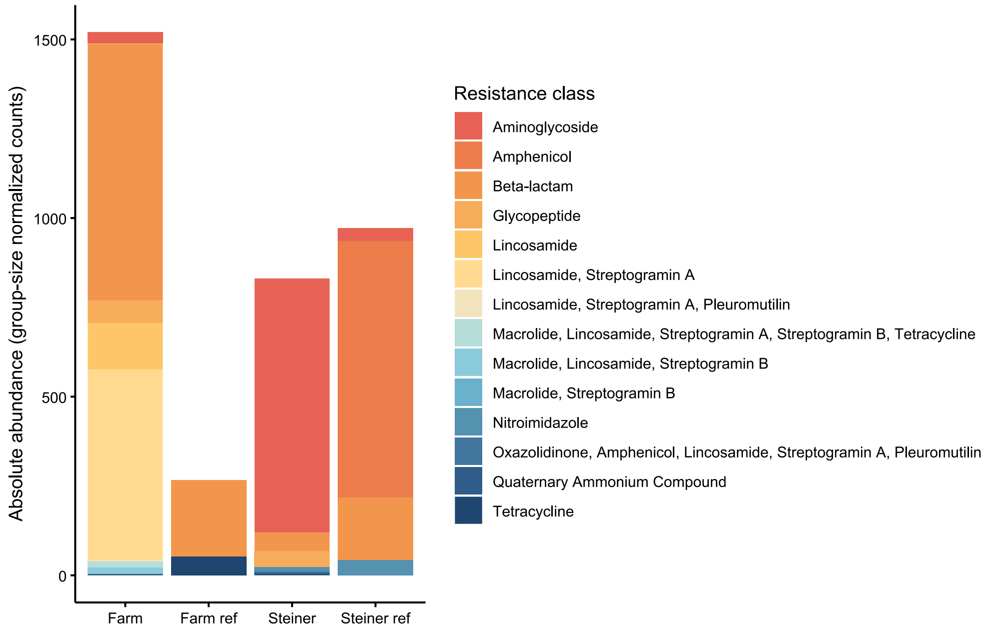


**Supplementary Figure 3 | Composition of group-exclusive antibiotic resistance genes.** Shown are the group-size normalized total abundances of lifestyle group-exclusive ARG classes. ARG counts were additionally normalized to the group size before the scaling step described under "Data normalization" in the main text. Farm: Farm group (N = 37), Farm ref: Farm reference group (N = 11), Steiner: Steiner group (N = 19), Steiner ref: Steiner reference group (N = 7).

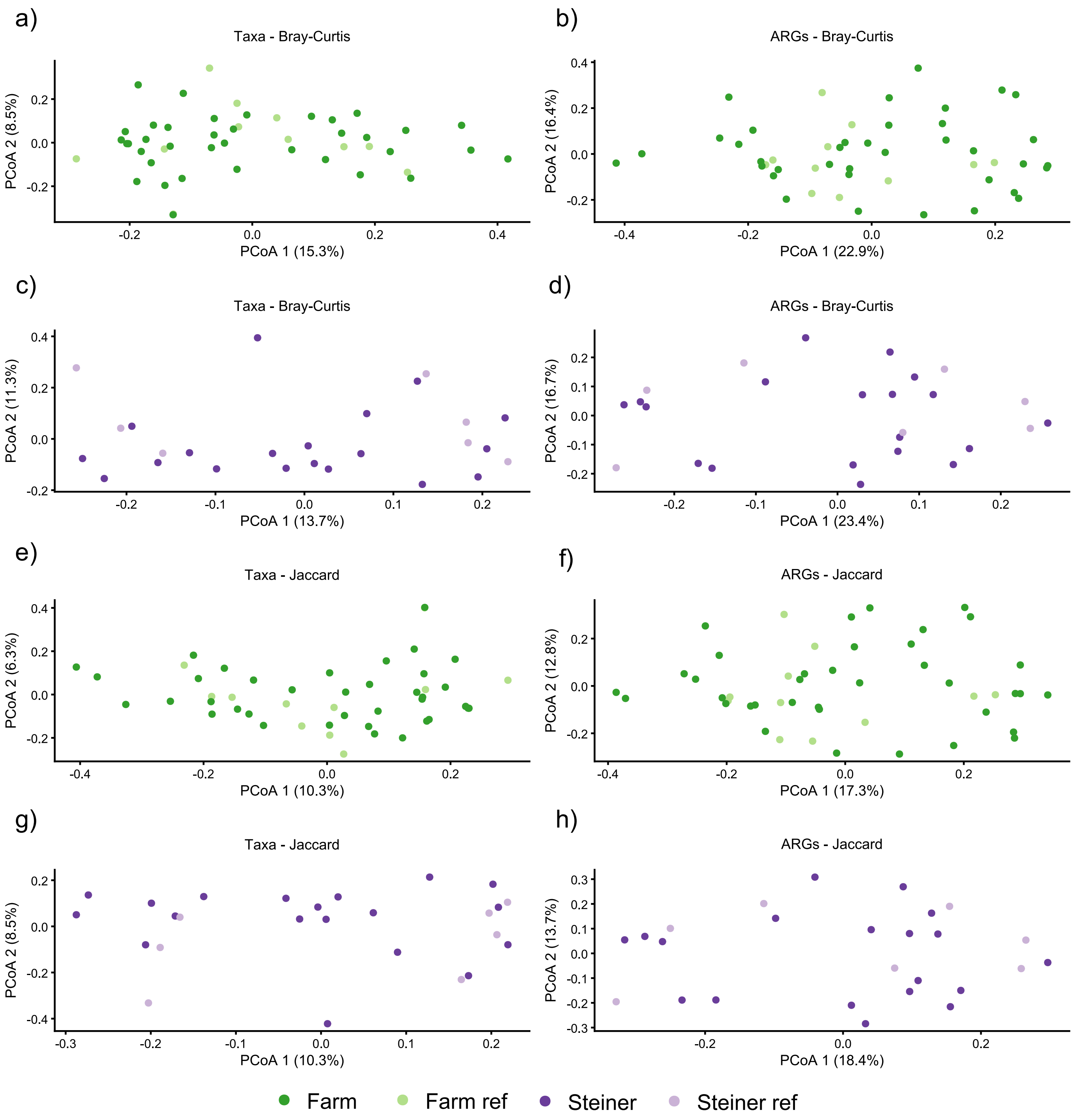


**Supplementary Figure 4 | Principal Coordinates Analyses of microbiomes and resistomes.** PCoA using either Bray-Curtis dissimilarity or Jaccard's similarity of **a** taxa (Farm and Farm ref), **b** ARGs (Farm and Farm ref), **c** taxa (Steiner and Steiner ref), **d** ARGs (Steiner and Steiner ref), **e** taxa (Farm and Farm ref), **f** ARGs (Farm and Farm ref), **g** taxa (Steiner and Steiner ref), **h** ARGs (Steiner and Steiner ref). Farm: Farm group (N = 37), Farm ref: Farm reference group (N = 11), Steiner: Steiner group (N = 19), Steiner ref: Steiner reference group (N = 7).

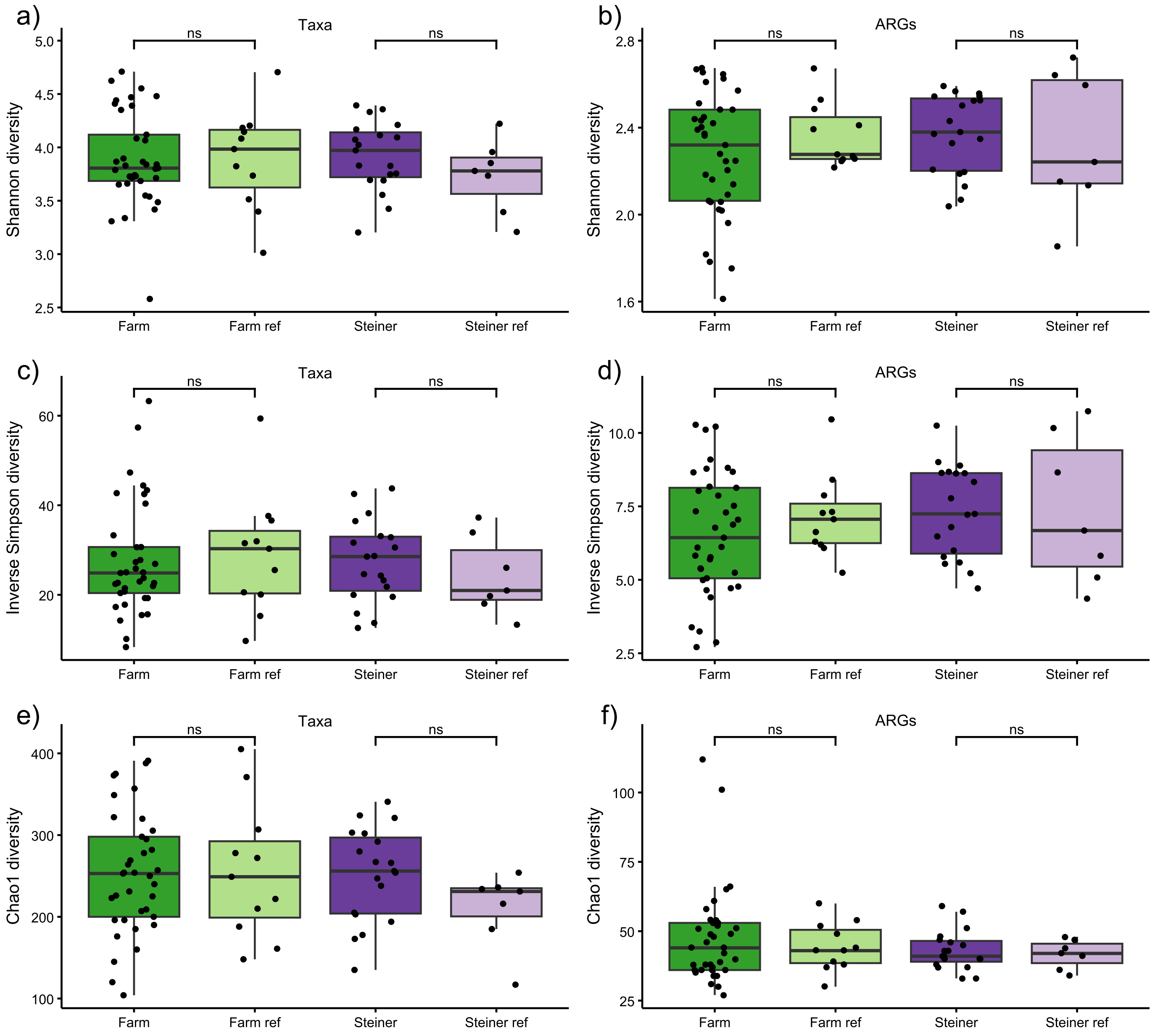


**Supplementary Figure 5 | Alpha diversity comparisons between groups.** Shannon, inverse Simpson and Chao1 alpha diversity indices of samples in different lifestyle groups. **a** Shannon diversity of bacterial species. **b** Shannon diversity of ARGs. **c** Inverse Simpson diversity of bacterial species. **d** Inverse Simpson diversity of ARGs. **e** Chao1 diversity of bacterial species. **f** Chao1 diversity of ARGs. Boxplot hinges represent 25 % and 75 % percentiles, the centre line represents the median. Length of whiskers denote 1.5 × IQR. The Mann–Whitney U test was used to compare diversity indices between the lifestyle groups and references, all comparisons were non-significant (ns, $p > 0.05$). Farm: Farm group (N = 37), Farm ref: Farm reference group (N = 11), Steiner: Steiner group (N = 19), Steiner ref: Steiner reference group (N = 7).

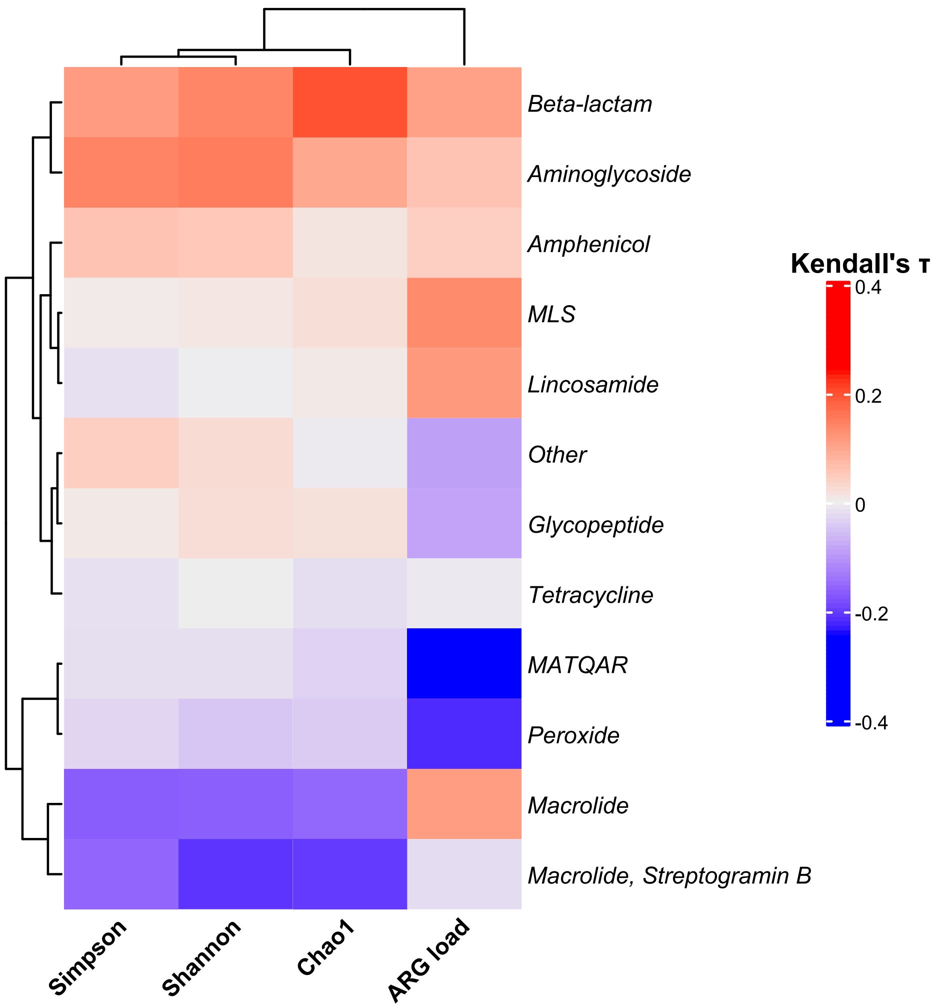


**Supplementary Figure 6 | Correlation of ARG classes with ARG load and inverse Simpson, Shannon and Chao1 diversities.** CLR transformed ARG class abundances were correlated with diversity measures using the Kendall rank correlation coefficient (τ). No correlations were significant after BH correction for multiple testing. Dendrograms are the result of row- and column-wise hierarchical clustering (complete-linkage) using Euclidian distances. Classes with a prevalence < 50% were aggregated to ”Other”. MLS: Macrolide, Lincosamide, Streptogramin B; MATQAR: Macrolide, Aminoglycoside, Tetracycline, Quinolone, Amphenicol, Rifamycin.
